# Prospective In Silico Simulation of the VESALIUS-CV Trial Using Biomedical Knowledge Graph and Real-World Data-Driven AI Modeling

**DOI:** 10.64898/2026.08.26.26361436

**Authors:** Amichai Perlman, Nir Goldstein, Marina Goldman, Michael Shapiro, Eran Barash, Alon Bar, Tamir Raveh, Eden Tordjman, Hallel Schussheim, Flavio Dormont, Omri Matalon

**Author notes:** Corresponding author: Omri Matalon, QuantHealth Ltd., Derech Begin 132, Tel Aviv, Israel. Prior presentation: This work was partly presented at the QuantHealth Clinical Development Summit, New York, NY, on May 27^th^ 2026. A preprint version of this manuscript was posted on medRxiv on August 26, 2026.

## Abstract

Cardiovascular outcomes trials are lengthy and costly. In silico trial simulation using real-world data may support earlier decision-making, but evidence generated before trial results are disclosed remains limited. We applied a semi-mechanistic machine-learning framework integrating patient-level real-world data with knowledge graph derived drug representations to simulate the VESALIUS-CV trial of evolocumab versus placebo before its results were public, using a locked model and prespecified trial design aligned with the trial protocol. Patient-level time-to-event predictions were aggregated into virtual trial arms to estimate cumulative incidence, hazard ratios (HR), confidence intervals (CI), and p-values for three-point major adverse cardiovascular events. In retrospective evaluation of the model’s patient-level predictive performance, time-dependent areas under the receiver operating characteristic curve ranged from 0.80 to 0.90 across follow-up horizons. In evaluation of retrospective trial-level predictive performance across 22 cardiovascular trials, the model predicted trial meeting the primary outcome with an accuracy of 0.91 (95% CI 0.77-1) and F1 score of 0.95 (95%CI 0.86-1). The prospective simulation predicted a significant reduction in major adverse cardiovascular events with evolocumab (HR 0.78; 95%CI, 0.70 to 0.87) at 54 months. The trial subsequently reported a HR of 0.75 (95% confidence interval, 0.65 to 0.86) at a median follow-up of 55 months, with progressive separation of event curves in both the simulation and the trial. The prospective simulation was concordant with the direction, magnitude, and temporal dynamics of the observed treatment effect, supporting in silico simulation as a tool for early decision-making, trial design, and de-risking in cardiovascular drug development.

## INTRODUCTION

Cardiovascular outcomes trials (CVOTs) represent one of the most resource-intensive components of clinical drug development. They are designed to evaluate the effect of interventions on major adverse cardiovascular events and are typically powered to detect relatively modest treatment effects. As a result, they require large patient populations and prolonged follow-up to accrue a sufficient number of events. CVOTs commonly enroll tens of thousands of participants, with estimated median costs per drug of approximately $141 million and per-patient costs approaching $35,000^1^. Collectively, large sample sizes, extended follow-up, and intensive operational requirements make CVOTs a high-cost, high-risk component of late-stage development and a major contributor to the overall financial burden of bringing new cardiovascular therapies to market^2^.

Across this development lifecycle, uncertainty regarding treatment effect remains substantial. Empirically, overall clinical success rates from early clinical development remain low, typically on the order of 10-30%^3,4^. In some analyses, the likelihood of approval for cardiovascular drugs from phase 1 has been estimated at below 10%^4^. In this environment, sponsors must make repeated, high-stakes decisions regarding asset and indication prioritization, sequencing of development programs, trial design choices, and continuation or termination of investment, often well before definitive randomized evidence becomes available.

Conventional planning approaches, such as feasibility assessments, historical benchmarks, and power calculations, provide useful structural guidance but are largely grounded in aggregate assumptions derived from prior trials or epidemiologic averages. However, treatment effects are inherently heterogeneous across patient populations due to variation in baseline risk and other covariates, implying that average effects may not generalize to specific subgroups. Indeed, when baseline risk varies, treatment effect heterogeneity will arise on at least one scale even within randomized studies^5^. Consequently, reliance on population-average assumptions offers limited insight into the expected magnitude, timing, and consistency of treatment effects across heterogeneous patient populations and evolving standards of care, leaving many strategic decisions insufficiently de-risked.

In this context, in silico trial simulation is increasingly viewed as a tool for continuous de-risking throughout drug development^6–8^. By enabling patient-level estimation of treatment effects under varying assumptions, indications, and patient subgroups, simulation has the potential to inform program prioritization, guide trial design and endpoint strategy, and support go/no-go decisions at multiple stages of development. Importantly, such insights are most valuable when they extend beyond trial sizing to address broader questions of expected clinical benefit, robustness across populations, and the timing of treatment effect emergence.

The growing availability of large-scale real-world data (RWD), together with advances in computational modeling, has accelerated interest in in silico trials as a complement to randomized evidence generation^9,10^. Rather than replacing randomized controlled trials, these approaches aim to provide earlier, population-level insight by emulating trial protocols within representative real-world cohorts. A central promise of this paradigm is the ability to prospectively estimate treatment effects across therapies and clinically relevant subgroups, thereby supporting trial de-risking and more informed decision-making prior to trial readout. However, empirical demonstrations of prospective predictive validity, in which simulations are conducted before results are known, remain limited, particularly for large cardiovascular outcomes trials. Addressing this gap is essential to establishing confidence in the broader use of in silico simulation across the development lifecycle.

### Modeling paradigms for cardiovascular risk prediction

Two broad paradigms dominate contemporary approaches to predicting clinical trial outcomes: mechanistic disease modeling^7,11^ and observational modeling grounded in RWD^12–14^.

Knowledge-based mechanistic models, often implemented as quantitative systems pharmacology (QSP) frameworks, encode biological hypotheses as coupled dynamical systems, typically ordinary differential equations, intended to represent causal relationships among biomarkers, disease processes, and drug mechanisms^15–17^. These models aim to simulate disease progression and treatment effects through explicit representation of intermediate biological states (e.g., lipid levels, plaque burden) and their evolution over time.

Mechanistic approaches offer several strengths for long-horizon cardiovascular outcomes prediction, including structured temporal extrapolation and the ability to simulate counterfactual scenarios under alternative biological assumptions. However, a central limitation lies in their capacity to adequately capture the heterogeneity of patient response observed in real-world populations. These models rely on predefined structures and parameterizations, often calibrated to aggregate data or limited experimental settings. This leads to limited representation of relevant patient features, variability in baseline risk, comorbidities, background treatments, and interaction effects across diverse patient subgroups.

A second paradigm is trial emulation based on RWD, which seeks to reproduce aspects of randomized trials by applying eligibility criteria, treatment definitions, and outcome ascertainment to observational healthcare data^12,18,19^. This approach leverages large-scale longitudinal RWD to estimate treatment effects directly from observed clinical practice, often using causal inference or survival analysis techniques. Its primary advantage is tight grounding in observed data, allowing capture of real-world heterogeneity in patient characteristics, background therapy, adherence, and care pathways^13^. However, trial emulation is inherently limited to what is observed in the data: it cannot reliably extrapolate beyond available follow-up, indications, or treatment contexts, and remains sensitive to residual confounding, selection bias, and data incompleteness. As a result, its utility for prospective prediction, particularly for long-horizon outcomes or novel trial settings, is constrained.

In this study, we describe the prospective simulation of the VESALIUS-CV trial using a hybrid position between these paradigms, combining large-scale RWD with structured biomedical knowledge to support population-level trial simulation. Rather than relying solely on direct trial emulation or explicit mechanistic equations, this framework integrates longitudinal clinical histories with drug-centric knowledge graph representations, enabling learning of treatment-outcome associations that are informed by both empirical data and curated biological structure. This semi-mechanistic design allows the model to move beyond pure observational replication while avoiding claims of explicit physiological causality^20^.

### Rationale for prospective simulation

A central challenge in evaluating in silico trial methodologies is distinguishing genuine predictive capability from post hoc model fitting. Many published demonstrations of trial simulation rely on retrospective benchmarking, in which models are developed, calibrated, or refined using train/test splits on data from trials whose outcomes are already known. While such analyses are valuable for assessing internal consistency and historical performance, they cannot fully address the question most relevant to clinical development decision-making: whether a model can generate reliable predictions *before* trial outcomes are available.

Prospective simulation requires strict temporal separation between model prediction and outcome disclosure. Model structure, parameters, eligibility criteria, endpoints, and analysis plans must be fixed prior to trial readout, and no information from interim analyses or unpublished results may inform the simulation. This separation is essential to avoid implicit leakage of outcome knowledge and to ensure that agreement between simulated and observed results reflects predictive signal rather than retrospective alignment. Without this discipline, apparent accuracy may overstate real-world utility. One rigorous approach to satisfying these requirements is to generate trial predictions prospectively, prior to trial completion or public readout, and to lock the simulation outputs until the actual trial results are published. Predicted and observed outcomes can then be compared. This prospective prediction framework was the approach used in the current study: all clinical trial simulation analyses were conducted well ahead of the study readout and locked in a publicly accessible, time-stamped record, preventing any modification of the simulation results^21^.

### Study objective

The objective of this study was to assess the agreement between a prespecified, locked in silico simulation and the direction, magnitude, and timing of treatment effects subsequently observed in the VESALIUS-CV randomized outcomes trial.

## METHODS

### Modeling framework

The modeling framework is a semi-mechanistic machine-learning approach designed to simulate population-level outcomes of randomized clinical trials using models trained on real-world longitudinal patient-level data combined with drug embeddings derived from a biomedical knowledge graph (**Figure 1**).

**Figure 1.**
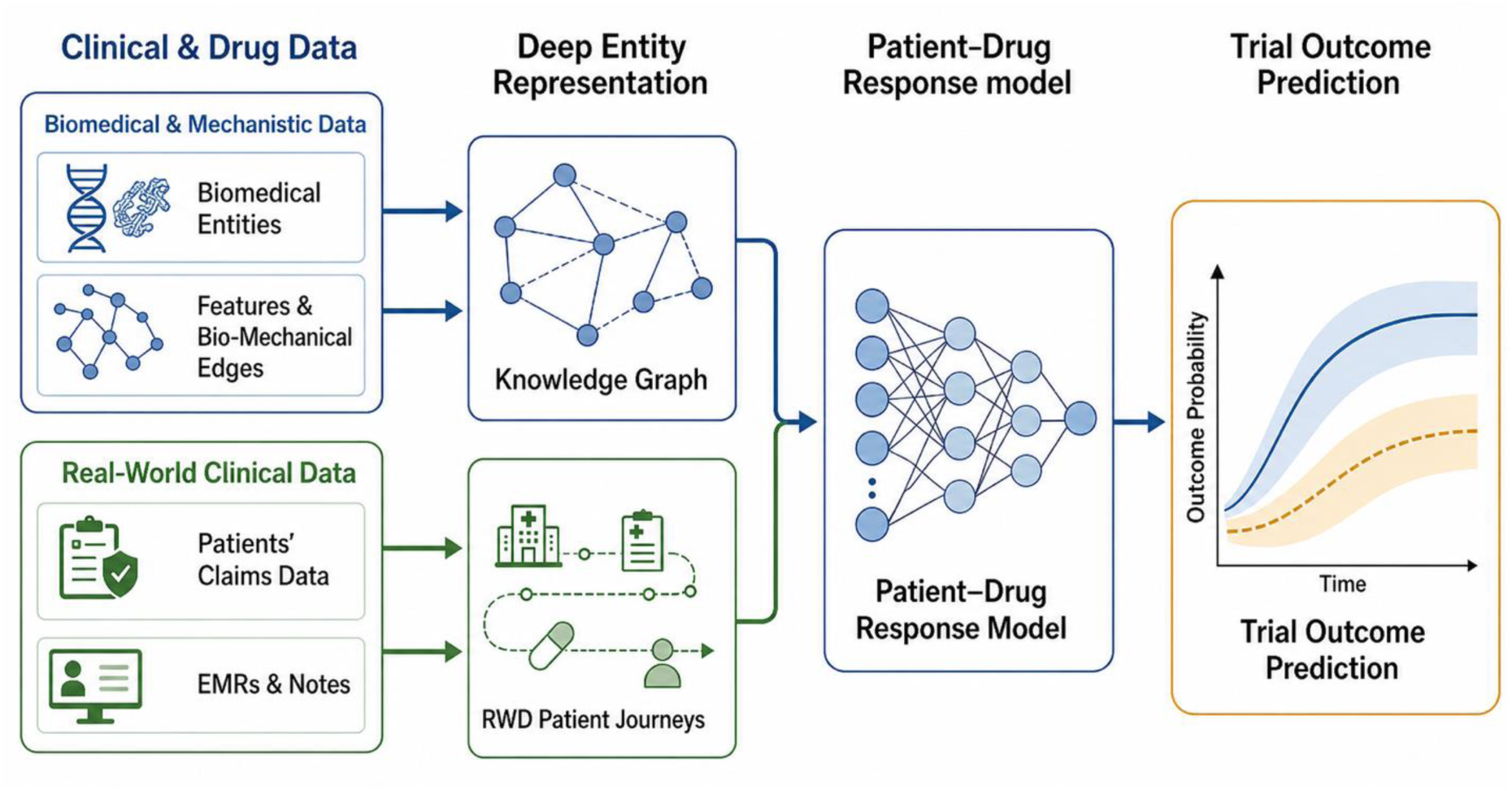
Predictive modeling framework and prospective trial simulation workflow. Biomedical knowledge and longitudinal real-world clinical data were integrated to construct patient-specific representations of disease biology and treatment response. Biomedical entities and their relationships were encoded through a knowledge graph capturing molecular, biological, and clinical associations, while patient-level information was derived from linked real-world data sources, including claims, electronic medical records, and treatment histories. These complementary data modalities were combined within a patient–drug response model to estimate individualized time-to-event outcomes. The trained model was subsequently applied prospectively to emulate the VESALIUS-CV trial population according to protocol-defined eligibility criteria. Treatment-specific outcome trajectories were generated for each simulated patient and aggregated to estimate trial-level efficacy endpoints, including survival curves, event rates, hazard ratios, and subgroup-specific treatment effects.

The framework learns associations between patient characteristics, treatment exposures, and time-to-event outcomes across heterogeneous real-world populations, while incorporating structured biological priors through knowledge graph embeddings that encode relationships between drugs, targets, pathways, and disease biology. This hybrid formulation introduces a latent mechanistic structure without requiring explicit systems-level equations and supports (i) modeling of real-world clinical heterogeneity and endpoint dynamics, consistent with clinical trial emulation principles, and (ii) generalization to previously unseen drugs by embedding them within the same biologically informed latent space. In the present study, patient-level time-to-event models estimated individualized risk trajectories for cardiovascular outcomes from baseline demographic and clinical features, longitudinal health histories, and treatment assignment. Additional details are provided in the **Supplementary Methods**.

### Data sources and ethics

Model development and trial simulation used two complementary real-world data sources covering the United States: longitudinal open claims data from PurpleLab^22^ and electronic health record (EHR) data from OMNY Health^23^, with patients de-identified and linked through a third-party tokenization service (Datavant Inc., San Francisco, CA, USA). Both sources provide longitudinal follow-up between 2014 and 2025 and include diagnoses, procedures, medication exposures, and demographic information; the EHR data additionally provide laboratory results and physiological measurements. All source data were standardized to the Observational Medical Outcomes Partnership (OMOP) Common Data Model^24,25^ to ensure consistent representation of clinical concepts and reproducible cohort definition (**Supplementary Methods**).

Drug exposures were represented using embeddings derived from a drug-centric biomedical knowledge graph of approximately 150,000 entities and more than 7 million relationships, constructed from over 30 curated databases, including DrugBank^26^, ChEMBL^27^, UniProt^28^, and Reactome^29^, and enriched through literature extraction with expert curation (**Supplementary Methods**).

This study used retrospective, de-identified patient-level data with no direct patient identifiers available to the researchers. As a retrospective analysis of de-identified data, the study received a determination of exemption from WCG IRB under 45 CFR § 46.104(d)(4) and was conducted in accordance with relevant guidelines and regulations. The simulated trial, VESALIUS-CV, is registered at ClinicalTrials.gov (NCT03872401); the authors did not conduct or have access to the trial.

### Outcome definition and follow-up

Cardiovascular outcomes were defined using prespecified code sets aligned with the VESALIUS-CV primary endpoint of 3-point major adverse cardiovascular events (MACE), comprising cardiovascular death, non-fatal myocardial infarction, and non-fatal stroke^30^. Time to event was measured from the index date to the first occurrence of the outcome or the end of available follow-up.

### Outcome modeling

Time to first event was modeled using ClinBoost, a patient-level survival modeling framework described previously^20^. Each patient is assigned an index date representing trial entry, and the outcome label captures both event occurrence and time to event or censoring, enabling appropriate handling of right-censored data. Patient representation incorporates structured clinical history summarized over predefined windows preceding the index date and treatment encoded from the unique drugs received within a fixed pre-index window. ClinBoost combines gradient-boosted trees^31^ with time-binned logistic regression layers^32^ to estimate event probabilities across discrete time intervals; these interval-specific predictions are transformed into individualized survival curves that form the basis for cumulative incidence estimation and trial simulation (**Supplementary Methods**).

### Prevention of information leakage

No data from VESALIUS-CV, including interim analyses, outcome estimates, or unpublished results, were used for model training, feature selection, parameter tuning, or validation. Model components, feature definitions, temporal windows, and analysis procedures were fixed before the prospective simulation^33^, and the simulation outputs were deposited in a time-stamped public record before trial readout^21^. For the primary analysis, no recalibration or adjustment was performed after trial results became available; performance was assessed solely by comparing the prespecified simulation outputs with the subsequently published trial outcomes.

### Virtual trial construction

Eligible patients were selected from the real-world data using prespecified criteria aligned with the VESALIUS-CV protocol. The cohort was defined to approximate a high-risk primary prevention population with enriched cardiometabolic and atherosclerotic burden: inclusion required evidence of at least one major cardiometabolic or vascular condition at baseline (type 2 diabetes mellitus, peripheral vascular disease, established atherosclerotic cardiovascular disease, or ischemic cerebrovascular disease) together with elevated low-density lipoprotein (LDL) cholesterol (≥90 mg/dL) or non-high-density lipoprotein (non-HDL) cholesterol (≥120 mg/dL) within the 6 months before cohort entry, and exclusion criteria removed patients with recent or unstable cardiovascular conditions and severe comorbidities (full criteria in the **Supplementary Methods**). For each patient, outcome trajectories were simulated under both the investigational and comparator treatment scenarios, generating patient-specific counterfactual predictions for each arm independent of actual treatment exposure. Randomization ratio, per-arm sample size, and follow-up duration matched those of the target trial. Patient-level predicted risk trajectories were aggregated within each arm to generate arm-level cumulative incidence functions, and relative treatment effects were summarized as hazard ratios. Event-risk trajectories were estimated using all eligible patients. These estimates were then used to calculate hazard ratios, 95% CIs, and P values, according to a standard analysis plan, assuming the target trial sample size of 6,150 patients per arm. (**Supplementary Methods**).

### Model validation

The cardiovascular outcomes model was evaluated, independently of the VESALIUS-CV simulation, at the patient level and at the trial level across past randomized cardiovascular outcomes trials with MACE as a primary or secondary endpoint, spanning diverse trial designs, therapeutic mechanisms, and baseline risk profiles. At the patient level, the real-world dataset was randomly partitioned into training (80%), validation (10%), and held-out test (10%) sets at the patient level, and time-to-event predictions were assessed on the test set using time-dependent receiver operating characteristic area under the curve (ROC-AUC) computed across discrete time bins, characterizing discrimination along the risk trajectory^34,35^. At the trial level, simulated outcomes were compared with published randomized trial results (**Table S1**) for agreement in the statistical success or failure of the investigational arm, in terms of meeting the trial defined primary outcome. Accuracy metrics with 95% confidence intervals were estimated by trial-level bootstrap (10,000 resamples). The proportion of trials without a statistically significant difference between simulated and observed HR was estimated using a log-scale normal approximation based on the reported confidence limits. Agreement between simulated and observed hazard ratios was additionally examined by Bland-Altman analysis (**Figure S1**).

### Concordance of the prospective simulation with observed results

Following readout, concordance between the prospective simulation and the reported VESALIUS-CV results was assessed across three prespecified dimensions. First, we assessed whether the simulation correctly predicted trial success, defined as a statistically significant treatment benefit on the primary outcome (hazard ratio <1 with P <0.05). Second, we evaluated the accuracy of the predicted treatment effect by comparing the predicted with the observed hazard ratio. Third, we compared arm-level event rates using the Kaplan–Meier estimate of the percentage of participants with an event at the final assessment.

### Post hoc population-matched sensitivity analysis

To assess the influence of population composition on simulated outcomes, the simulation was repeated in a cohort constructed to match the baseline characteristics reported for the trial population, including demographics (age, sex, race, and ethnicity) and key clinical variables (hypertension, obesity, and diabetes). Race and ethnicity were taken as recorded in the source claims and EHR data, using the fixed categories of those sources. Results were compared with the primary simulation to assess the sensitivity of relative and absolute treatment effect estimates to population composition (**Supplementary Methods**).

## RESULTS

### Summary of model validation

A patient-level survival model predicting time to first 3-point MACE was developed using longitudinal clinical histories and treatment representations derived from biologically informed drug embeddings, trained on a real-world longitudinal dataset of 868,533 patients.

On the held-out test set, time-dependent ROC-AUC values across discrete time bins spanning up to approximately 6.5 years of follow-up were 0.90 at ≤11 days, 0.86 at ≤120 days, and approximately 0.81–0.82 at longer-term horizons extending beyond 5 years (**Figure 2**). While a modest decline in AUC was observed over time, performance remained stable, providing robust discrimination for both short-term and long-term cardiovascular risk.

**Figure 2.**
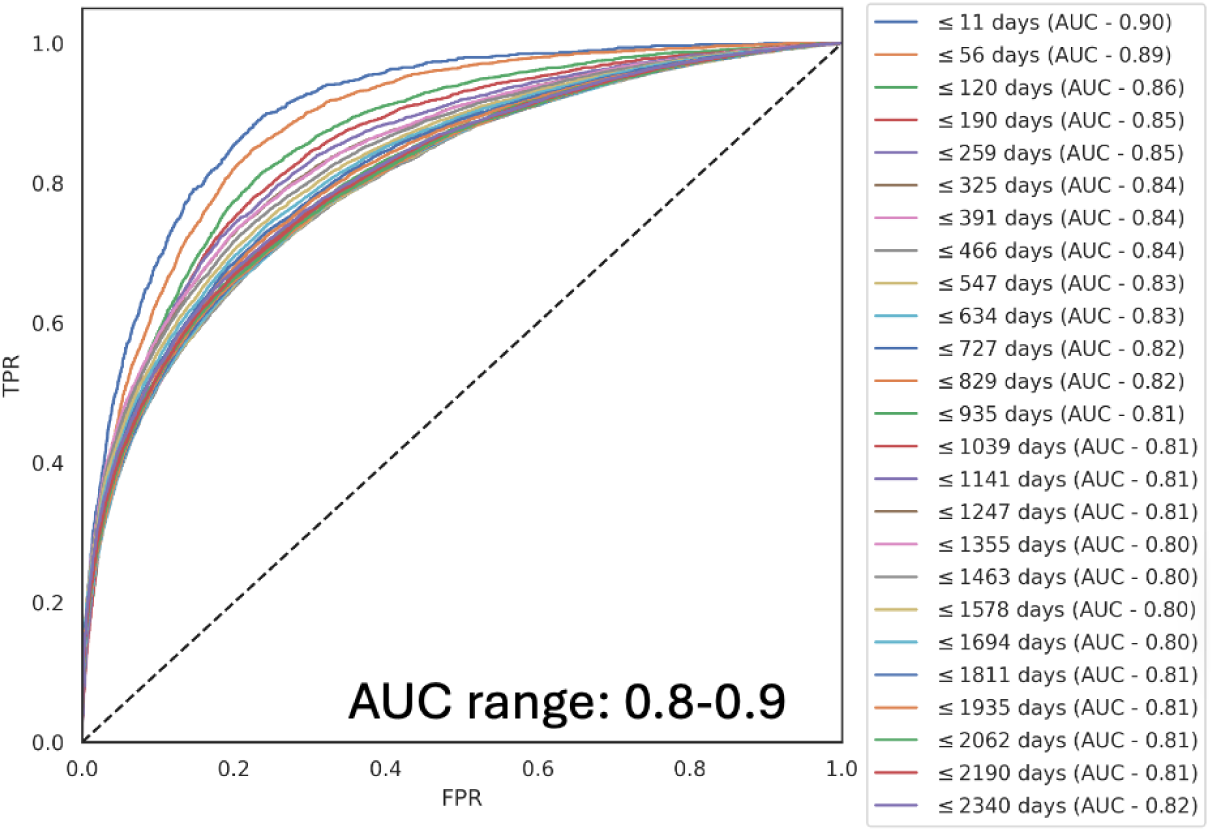
Model validation performance. Time-dependent receiver operating characteristic curves on the held-out real-world test set across follow-up horizons (area under the curve [AUC] range, 0.80–0.90).

Trial-level performance was validated against 22 large cardiovascular outcomes trials using 3-point MACE as the endpoint, spanning multiple therapeutic classes and mechanisms of action, including glucagon-like peptide-1 receptor agonists, sodium–glucose cotransporter-2 inhibitors, antiplatelet therapy, dipeptidyl peptidase-4 inhibitors, insulin analogs, and other cardiometabolic agents. (**Table S1**) Simulated hazard ratios showed good concordance with observed outcomes, with an F1 score of 0.95 (95%CI 0.86-1) and accuracy of 0.91 (95% CI 0.77-1); the Bland–Altman mean bias between observed and simulated hazard ratios was -0.09 (limits of agreement, -0.32 to 0.14) (**Figure S1**), and 18 of 22 trials (81.8%) had a predicted HR not significantly different from the observed HR.

### Prospective simulation of VESALIUS-CV

The simulation of VESALIUS-CV was conducted prior to public disclosure of the trial results, with model structure, parameters, eligibility criteria, endpoints, and analysis procedures locked in advance^21^. The simulation preceded publication of the trial results and was completed without access to interim analyses or any unpublished outcome data. Eligibility criteria were aligned with the VESALIUS-CV protocol, and the primary simulated endpoint was 3-point MACE (cardiovascular death, myocardial infarction, or ischemic stroke), consistent with the trial’s primary efficacy endpoint. Model outputs were generated at prespecified follow-up horizons to reflect both intermediate and longer-term trial readouts.

The simulation cohort, defined from the public protocol^36^, represents a high-risk cardiometabolic population with a high prevalence of type 2 diabetes, vascular disease, and elevated lipid levels, consistent with populations targeted in contemporary cardiovascular outcomes trials. The eligible cohort comprised 39,777 patients, with a mean age of 68.5 years (median 69 years) and a predominance of female patients (61.2%), and 6,150 patients were simulated per arm. The population reflects an older, high-risk group with substantial underlying cardiometabolic burden, appropriate for evaluating cardiovascular outcomes in a trial-like setting. Baseline characteristics of the simulation cohort and of the trial population are provided in **Table 1**.

**Table 1.** Baseline characteristics of the simulation cohort and of the VESALIUS-CV trial population.

| Characteristic | Simulation cohort<br>(N = 39,777) | VESALIUS-CV<br>evolocumab<br>(N = 6,129) | VESALIUS-CV<br>placebo<br>(N = 6,128) |
|---|---|---|---|
| **Demographics** |  |  |  |
| Age, median (IQR), years | 69 (63–74) | 66 (60–71) | 66 (60–71) |
| Female sex, no. (%) | 24,326 (61.2) | 2,619 (43) | 2,595 (42) |
| Race, no. (%)<sup>a</sup> |  |  |  |
| White | 28,538 (71.7) | 5,708 (93) | 5,693 (93) |
| Black or African American | 8,001 (20.1) | — | — |
| Asian | 801 (2.0) | — | — |
| Hispanic or Latino ethnicity, no. (%)<sup>a</sup> | 4,329 (10.9) | 1,017 (17) | 1,016 (17) |
| BMI, median (IQR), kg/m<sup>2</sup> | 34 (29–40) | 30 (27–34) | 30 (27–33) |
| **Coexisting conditions, no. (%)** |  |  |  |
| Hypertension | 37,448 (94.1) | 5,351 (87) | 5,319 (87) |
| Diabetes | 34,497 (86.7) | 3,598 (59) | 3,524 (58) |

Trial values are as reported by Bohula et al.<sup>36</sup> Em dashes (—) denote categories not separately reported in the trial publication. BMI, body mass index; HDL, high-density lipoprotein; IQR, interquartile range; LDL, low-density lipoprotein. <sup>a</sup>Race and ethnicity as recorded in the source claims and electronic health record data (see Methods).

### Simulation results and comparison with observed trial results

After a median follow-up of 55 months, the VESALIUS-CV trial reported 5-year Kaplan–Meier estimates of 3-point MACE of 6.2% in the evolocumab group and 8.0% in the placebo group, corresponding to a hazard ratio of 0.75 (95% CI, 0.65–0.86; P <0.001)^36^. The prospective simulation generated prior to readout predicted treatment and control arm event rates and relative treatment effects at 54 months of follow-up. The simulation predicted a 3-point MACE event rate of 8.8% in the evolocumab arm and 11.2% in the placebo arm, corresponding to a simulated hazard ratio of 0.78 (95% CI, 0.70–0.87; P <0.01), demonstrating close agreement with the observed estimate, including overlapping confidence intervals and consistent directionality of effect (**Figure 3A,B** and **Table 2**).

**Figure 3.**
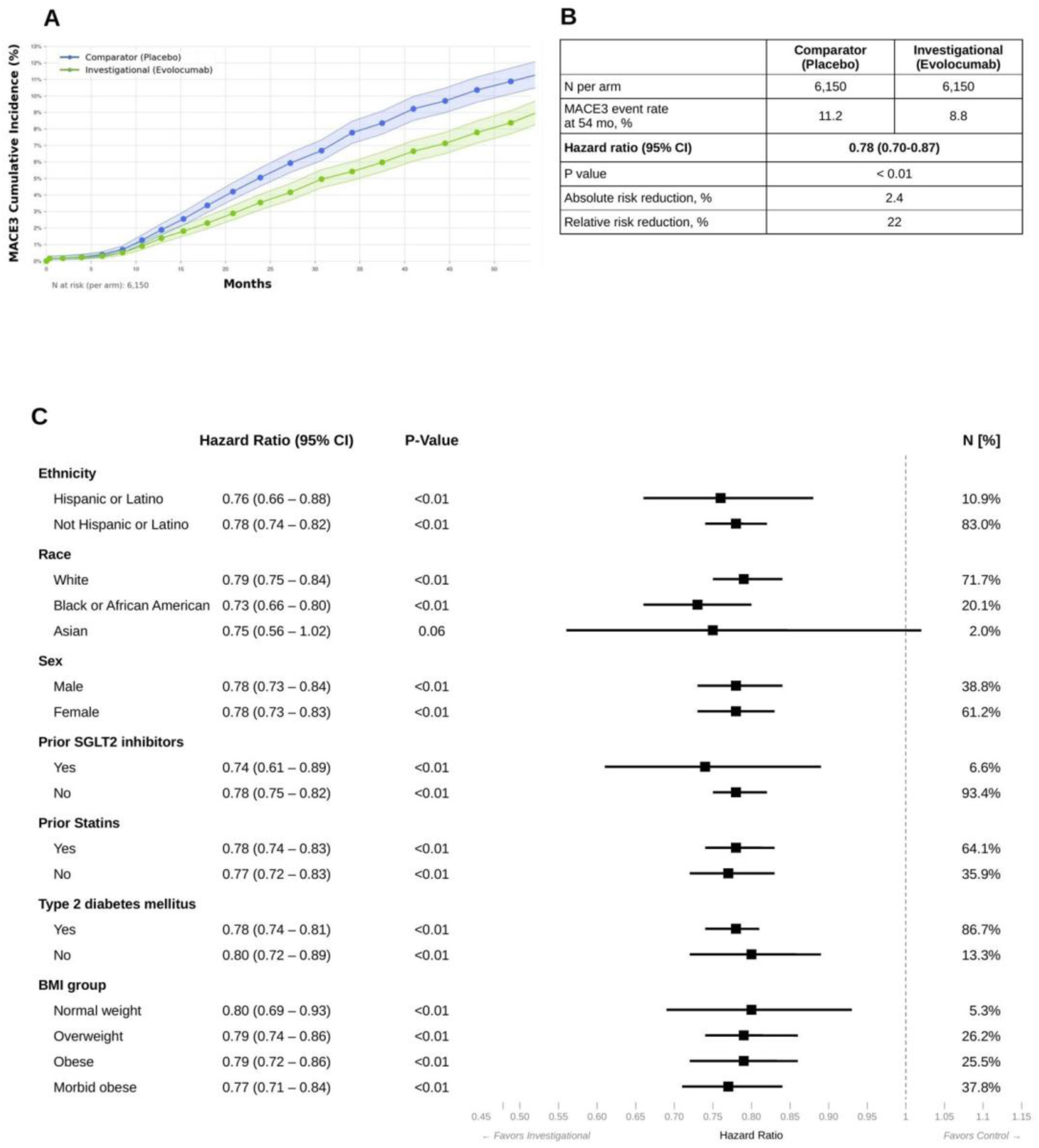
Prospective simulation of VESALIUS-CV. (A) Simulated cumulative incidence of 3-point major adverse cardiovascular events (MACE; labeled MACE3) for the comparator (placebo) and investigational (evolocumab) arms over 54 months, with 95% confidence intervals assuming 6,150 per arm. (B) Summary of simulation results at 54 months: event rates, hazard ratio, and absolute and relative risk reduction. (C) Simulated hazard ratios by baseline subgroup, with 95% confidence intervals and the proportion of the simulation cohort in each subgroup.

**Table 2.**
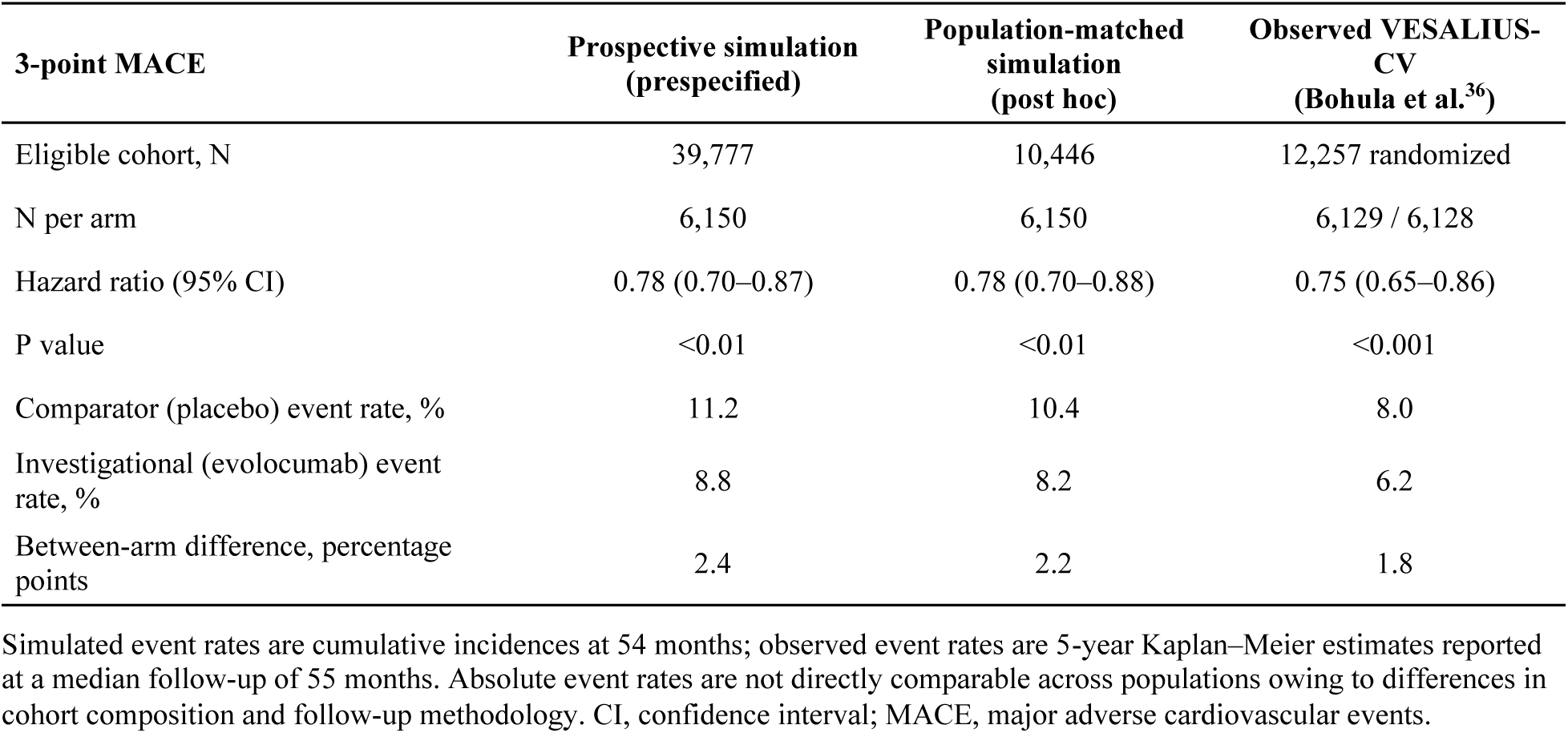
Observed VESALIUS-CV results compared with the prospective simulation and the post hoc population-matched simulation.

| **3-point MACE** | **Prospective simulation (prespecified)** | **Population-matched simulation (post hoc)** | **Observed VESALIUS-CV (Bohula et al.<sup>36</sup>)** |
|---|---|---|---|
| Eligible cohort, N | 39,777 | 10,446 | 12,257 randomized |
| N per arm | 6,150 | 6,150 | 6,129 / 6,128 |
| Hazard ratio (95% CI) | 0.78 (0.70–0.87) | 0.78 (0.70–0.88) | 0.75 (0.65–0.86) |
| P value | <0.01 | <0.01 | <0.001 |
| Comparator (placebo) event rate, % | 11.2 | 10.4 | 8.0 |
| Investigational (evolocumab) event rate, % | 8.8 | 8.2 | 6.2 |
| Between-arm difference, percentage points | 2.4 | 2.2 | 1.8 |

Simulated event rates are cumulative incidences at 54 months; observed event rates are 5-year Kaplan–Meier estimates reported at a median follow-up of 55 months. Absolute event rates are not directly comparable across populations owing to differences in cohort composition and follow-up methodology. CI, confidence interval; MACE, major adverse cardiovascular events.

Simulated cumulative incidence curves showed gradual divergence between treatment and control arms, with increasing separation over time. This pattern was concordant with the Kaplan-Meier curves observed in VESALIUS-CV^36^, which similarly exhibited progressive separation during follow-up, consistent with the delayed accrual of benefit characteristic of LDL cholesterol-lowering therapies.

Simulated subgroup analyses demonstrated that the predicted treatment effect was generally consistent across all examined patient subgroups, consistent with the observed results. Hazard ratios were uniformly below 1.0 across categories of age, sex, race, body mass index (BMI), and key clinical characteristics, indicating a consistent direction of benefit. The magnitude of effect was similar across most subgroups, with overlapping confidence intervals and no evidence of meaningful heterogeneity (**Figure 3C**). Overall, the simulation recapitulated the pattern observed in the VESALIUS-CV trial, supporting the robustness and generalizability of the predicted treatment effect across diverse patient populations.

### Post hoc analysis of residual differences

The simulated effect size, in terms of both the hazard ratio and the between-arm difference in event rates, was concordant with the observed effect; however, the absolute event rates in each arm were 2.6-3.2 percentage points higher in the simulation. This discrepancy may reflect differences between the simulated population and the population actually enrolled in the trial. To investigate this, a post hoc analysis was conducted in which the simulation was rerun using a population more closely aligned with the enrolled cohort, constructed to match the trial population across key characteristics, including demographics (age, sex, race, and ethnicity) and comorbidities (hypertension, obesity, and diabetes).

Following this population matching, the predicted treatment effect remained unchanged (hazard ratio 0.78; 95% CI, 0.70–0.88), while the discrepancy between the simulated and observed event rates was reduced, although a residual difference in absolute event rates per arm remained (2.0-2.4 percentage points) (**Figure 4A,B** and **Table 2**).

**Figure 4.**
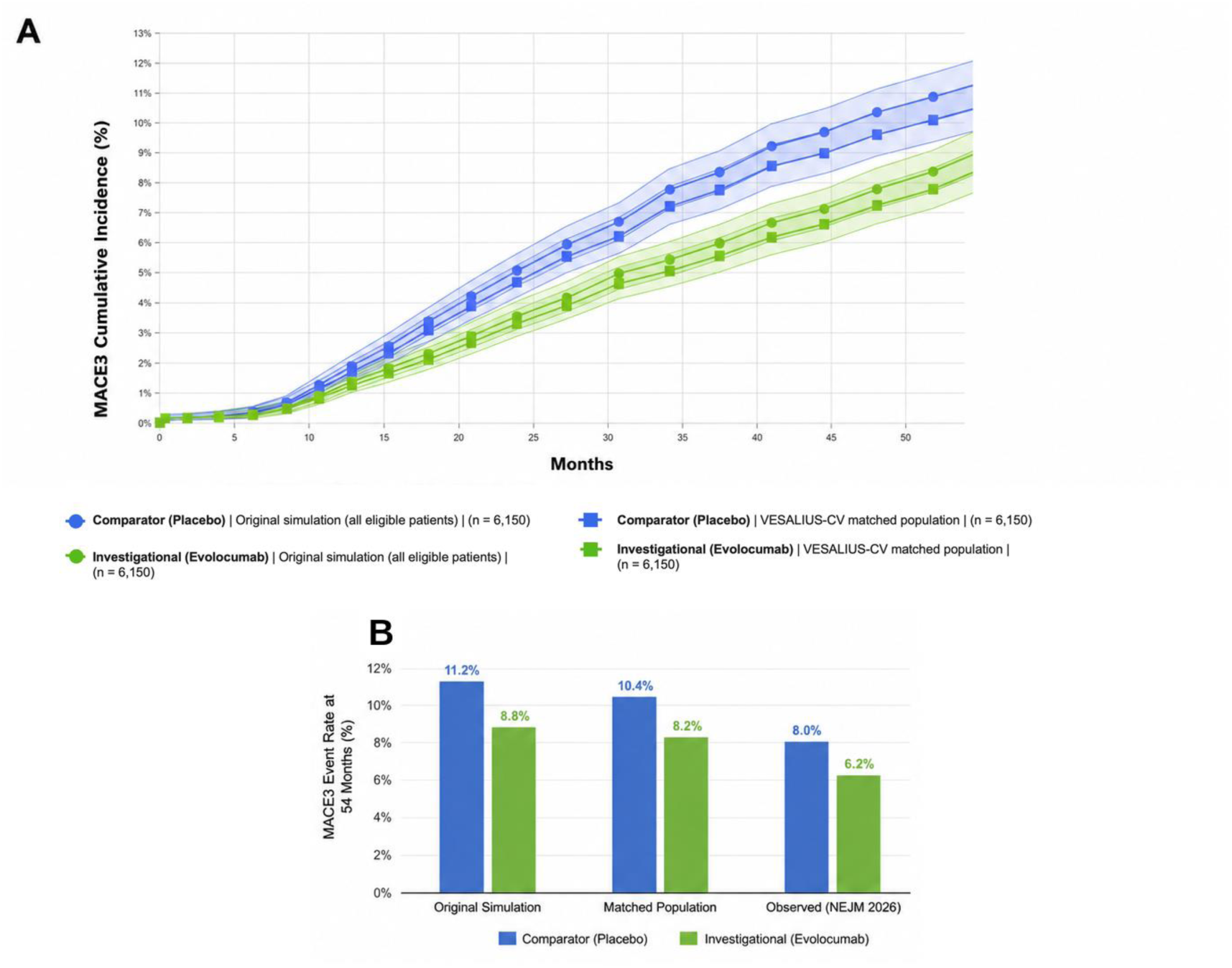
Post hoc population-matched sensitivity analysis. (A) Simulated cumulative incidence of 3-point major adverse cardiovascular events (MACE; labeled MACE3) over 54 months in the original simulation cohort (all eligible patients; circles) and in the cohort matched to the reported VESALIUS-CV baseline characteristics (N = 10,446; squares); n = 6,150 per arm in both cohorts; shaded regions denote 95% confidence intervals. (B) Event rates by arm at 54 months in the original and population-matched simulations compared with the observed 5-year Kaplan–Meier estimates reported for VESALIUS-CV, showing convergence toward the observed rates with population matching. Corresponding hazard ratios and event rates are given in Table 2.

## DISCUSSION

This study evaluates a machine learning–driven in silico trial simulation framework through patient-level predictive validation, retrospective trial-level benchmarking, and a fully prospective application to the VESALIUS-CV trial. At the patient level, the model demonstrated strong and durable discriminatory performance across time horizons, with ROC-AUC values ranging from 0.90 in the short term to approximately 0.81–0.82 over longer follow-up, supporting its ability to accurately capture individual cardiovascular risk trajectories. At the trial level, validation across 22 large cardiovascular outcomes trials showed good concordance between simulated and observed treatment effects across diverse therapeutic classes and study designs.

Building on this multi-level validation, the prospective simulation of VESALIUS-CV, conducted prior to public disclosure of trial results, accurately anticipated the direction, magnitude, and temporal evolution of treatment benefit observed in the randomized trial. In addition, subgroup analyses demonstrated consistent predicted treatment effects across a broad range of patient subpopulations, including stratifications by demographics and baseline clinical characteristics, with no evidence of meaningful heterogeneity. Together, these findings provide empirical support that a machine learning-based, RWD-driven framework can generate accurate and generalizable trial-level simulations of cardiovascular outcomes trials.

While differences in absolute event rates were observed, the simulation showed strong concordance with the observed VESALIUS-CV results for the relative treatment effect and close agreement for the between-arm difference in event rates. Simulated event rates exceeded those reported in the trial, with a between-arm difference of 2.4 percentage points compared with 1.8 percentage points in the observed results. Following post hoc alignment of key population characteristics, the gap between simulated and observed absolute event rates was reduced, and the simulated between-arm difference moved closer to the observed value (2.2 versus 1.8 percentage points).

This pattern highlights an important methodological distinction between relative and absolute measures of treatment effect. The close agreement in hazard ratios, together with the close approximation of the between-arm difference in event rates, despite residual differences in absolute event rates, suggests that the simulation framework captures the underlying treatment effect with high fidelity, even when baseline risk is not perfectly calibrated. This is consistent with prior evidence that relative treatment effects are generally more stable and transportable across populations than absolute risk estimates, which are more sensitive to differences in patient characteristics, event definitions, and measurement processes^37–39^. The remaining differences in absolute event rates may therefore reflect these sources of variability. More broadly, the ability to generate longitudinal trial simulations enables treatment effects to be evaluated across multiple complementary dimensions, including hazard ratios, cumulative event rates, event-rate differences between treatment arms, and the temporal evolution of treatment benefit, providing a richer basis for trial evaluation and design.

The partial attenuation of the discrepancy following population matching supports the interpretation that baseline risk calibration, rather than treatment effect estimation, is the primary driver of these differences. Taken together, these findings underscore the importance of a multi-dimensional evaluation framework for in silico trial performance, in which binary trial success, relative treatment effects, and absolute event rates are considered jointly. Within this framework, relative effect estimates may provide the most robust and decision-relevant signal for prospective applications, while absolute risk estimates remain essential for contextualizing clinical impact and informing trial design assumptions.

### Implications for cardiovascular drug development

The VESALIUS-CV simulation illustrates the framework’s ability to generalize treatment-effect predictions across distinct clinical settings. Although evolocumab had previously been evaluated in the FOURIER trial^40^, VESALIUS-CV enrolled a substantially different population, consisting of high-risk patients without prior myocardial infarction or stroke, with different baseline lipid profiles and longer follow-up. These differences introduce uncertainty to direct extrapolation of treatment effects from prior trials. Rather than relying on historical trial outcomes, the simulation estimated treatment effects under the specific eligibility criteria, population characteristics, and follow-up assumptions of the VESALIUS-CV protocol. The close agreement between predicted and observed outcomes suggests that integrating longitudinal real-world patient data with biologically informed drug representations may enable treatment-effect prediction across new trial settings and patient populations.

The ability to generate accurate predictions across distinct patient populations and trial designs has important implications for cardiovascular drug development. Prospective in silico trial simulation has the potential to support earlier and more informed decision-making in cardiovascular drug development. Such simulations can inform go/no-go decisions prior to trial initiation by providing quantitative expectations regarding the likelihood of observing a clinically meaningful effect. This capability is particularly valuable for large, long-duration cardiovascular outcomes trials, where late-stage failure carries substantial clinical and financial cost.

Beyond binary decisions, prospective simulations may help support trial design, including evaluating the potential impact of eligibility criteria and the enrolled population on treatment effect, estimation of trial size and the follow-up duration required to detect treatment effects, evaluation of endpoint selection, and assessment of the impact of baseline risk and background therapy on event accrual.

### Positioning relative to clinical trial emulation and mechanistic models

Clinical trial emulation (CTE) and QSP represent two established but distinct approaches to simulating cardiovascular trials. QSP models encode biological mechanisms and causal hypotheses across pathways, biomarkers, and disease progression, enabling mechanistic interpretation but depending heavily on prior knowledge and calibration data.

CTE approaches, by contrast, use observational clinical data to approximate randomized trial conditions, offering strong empirical validity, real-world generalizability, and the ability to capture patient heterogeneity and clinical endpoints. However, they are generally limited to drugs already observed in the data and lack explicit biological reasoning.

The framework described here can be viewed as a semi-mechanistic bridge between these paradigms. It combines longitudinal clinical data with knowledge graph embeddings that encode drug properties, mechanisms, and relationships, introducing a biologically structured prior into a data-driven trial simulation model.

This preserves key strengths of CTE, including modeling diverse patients and real-world endpoints, while addressing a major limitation: generalizing to previously unseen drugs. By representing drugs in a biologically informed embedding space, the model can simulate new interventions without requiring a fully specified systems-level QSP model.

Thus, this semi-mechanistic approach offers a practical and scalable alternative to purely mechanistic or purely observational methods, combining empirical performance, biological plausibility, and broader generalizability in cardiovascular trial simulation.

### Limitations and future directions

Several limitations should be considered when interpreting these findings. At the same time, many of these limitations represent active areas of methodological development and opportunities for future refinement.

First, the approach depends on the quality, completeness, and representativeness of real-world data. Observational datasets may differ systematically from randomized trial populations due to selection bias, missing data, and unmeasured confounding, which are not always fully captured by observed covariates. To mitigate these risks, our framework incorporates rigorous cohort construction and validation against epidemiological data and completed clinical trials. Ongoing improvements may include the integration of additional data sources (e.g., registries, omics, or imaging) and integration of explicit causal inference techniques.

Second, while the model integrates biologically informed representations through knowledge graph embeddings, it does not provide explicit, fully specified mechanistic causality in the way QSP models do. Future work under development includes enhancing explicit interpretability through hybridization with mechanistic sub-models, incorporation of causal graphs, and systematic post hoc model interrogation methods.

Third, the simulation inherently relies on assumptions regarding the characteristics of the trial to be emulated, including the patient population that will be enrolled, the rate and duration of enrollment, and the overall trial duration. In practice, these elements are uncertain at the design stage and may differ from realized trial execution. The framework addresses this by enabling flexible scenario analysis: investigators can vary inclusion criteria, enrollment dynamics, and follow-up duration to explore a range of plausible outcomes and sensitivities. However, predictions remain contingent on the specified assumptions, and inaccuracies in these inputs may impact simulated results. Future enhancements could include tighter integration with operational trial data and adaptive updating as enrollment progresses.

Finally, while the ability to simulate previously unseen drugs represents a key strength of the semi-mechanistic approach, it also introduces uncertainty when extrapolating beyond the observed data distribution. This risk is mitigated through biologically grounded embeddings and validation against known drug classes, but careful interpretation is required, particularly for novel mechanisms of action. Continued expansion of the underlying knowledge graph and incorporation of experimental or early-phase data may further improve robustness in these settings.

Beyond cardiovascular disease, the framework is designed as a general-purpose clinical trial simulation platform that can be applied across therapeutic areas wherever longitudinal patient data and clinically relevant outcomes are available. The underlying methodology is not indication-specific and has already been applied across more than 20 disease areas and clinical development programs, with ongoing expansion to additional indications, endpoints, and treatment modalities. Continued validation across diverse therapeutic settings will further characterize the scope and performance of the approach and help refine its application to an increasingly broad range of clinical development questions.

Taken together, the results of this study support the potential of RWD-driven, biologically informed simulation as a scalable decision-support tool for clinical development, capable of generating prospective insights to inform trial design, portfolio strategy, and evidence generation across a wide range of therapeutic contexts.

## STUDY HIGHLIGHTS

### What is the current knowledge on the topic?

In silico clinical trial simulation using real-world data and machine learning has shown retrospective agreement with completed trials, but prospective evidence, in which predictions are generated and locked before trial readout, remains scarce, particularly for large cardiovascular outcomes trials.

### What question did this study address?

Whether a prespecified, locked simulation of the VESALIUS-CV trial, produced by a semi-mechanistic model that combines longitudinal real-world data with knowledge graph–derived drug representations, agreed with the direction, magnitude, and timing of the treatment effect subsequently observed in the trial.

### What does this study add to our knowledge?

The prospective simulation predicted a hazard ratio of 0.78 (95% CI, 0.70–0.87) for 3-point MACE with evolocumab versus placebo; the trial reported 0.75 (95% CI, 0.65–0.86), with concordant progressive separation of event curves and consistent effects across subgroups. Relative treatment effects were well reproduced, whereas absolute event rates were sensitive to the composition of the simulated population.

### How might this change clinical pharmacology or translational science?

Prospective, time-stamped simulation provides a transparent standard for evaluating in silico trial methods and supports their use as complementary tools for go/no-go decisions, eligibility and endpoint design, and sample-size and follow-up planning in cardiovascular drug development.

## Data Availability

The datasets used in this study, including real-world data (RWD) derived from electronic medical records and insurance claims, as well as components of the biomedical knowledge graph, were obtained under license from proprietary vendors and cannot be made publicly available. Researchers interested in accessing the datasets may contact the corresponding author to discuss the possibility of data access, subject to licensing agreements and confidentiality obligations.

## ACKNOWLEDGEMENTS

Large language models (Claude by Anthropic and ChatGPT by OpenAI) were used as assistive tools for manuscript preparation. Figures presenting study results were generated directly from the underlying data using python and do not represent generative-model approximations. All analyses, figures, and manuscript content were reviewed, edited, and verified by the authors, who take full responsibility for the final manuscript.

## AUTHOR CONTRIBUTIONS

A.P., E.T., F.D., M.G., M.S., E.B., A.B., T.R., H.S., and O.M. wrote the manuscript; N.G., H.S., A.P., T.R., F.D., and O.M. designed the research; N.G., H.S., A.P., O.M., M.G., E.B., M.S., and A.B. performed the research; N.G., H.S., A.P., and O.M. analyzed the data.

## DATA AVAILABILITY STATEMENT

The patient-level claims and electronic health record data used in this study were obtained under license from PurpleLab and OMNY Health and cannot be shared publicly owing to contractual and privacy restrictions; access may be requested from the respective data providers. The biomedical knowledge graph and the ClinBoost modeling framework are proprietary to QuantHealth Ltd. The prespecified, time-stamped simulation outputs for VESALIUS-CV deposited before trial readout are publicly available on the Open Science Framework at https://osf.io/r6eqa Aggregate simulation results supporting the findings of this study are provided in the article and its Supplementary Material.

## SUPPLEMENTARY MATERIAL

Supplementary Methods. Modeling framework, data standardization, construction of longitudinal patient histories, ClinBoost architecture, simulation cohort eligibility criteria, generation of simulated trial outcomes, and post hoc population-matched analysis.

Table S1. Randomized cardiovascular outcomes trials used for trial-level model validation.

Figure S1. Bland–Altman analysis of observed versus simulated hazard ratios across 22 trials.

