## Supplementary for "Prospective In Silico Simulation of the VESALIUS-CV Trial Using Biomedical Knowledge Graph and Real-World Data-Driven AI Modeling"

**Contents**

Supplementary Methods

Table S1. Randomized cardiovascular outcomes trials used for trial-level model validation

Figure S1. Bland–Altman analysis of observed versus simulated hazard ratios

Supplementary References

### SUPPLEMENTARY METHODS

#### *S1. Modeling framework*

The framework leverages large-scale observational data to learn associations between patient characteristics, treatment exposures, and time-to-event clinical outcomes, as observed across heterogeneous real-world populations. In parallel, it incorporates structured biological priors through knowledge graph embeddings, which encode relationships between drugs, targets, pathways, and disease biology. This hybrid formulation allows the model to move beyond purely observational clinical trial emulation, introducing a latent mechanistic structure without requiring explicit systems-level equations.

In the current study, the framework involves models simulating patient-level time to event that estimate individualized risk trajectories for cardiovascular outcomes over time. These models integrate baseline demographic and clinical features, longitudinal health histories, and treatment assignments. Drug exposure is represented using multidimensional embeddings derived from a biomedical knowledge graph, capturing each therapy's relationships to biological and physiological pathways.

This representation enables two key capabilities: (i) accurate modeling of real-world clinical heterogeneity and endpoint dynamics, consistent with clinical trial emulation principles, and (ii) generalization to previously unseen drugs, by embedding them within the same biologically informed latent space. As a result, the framework supports simulation, prediction, and optimization of clinical trials under both observed and novel therapeutic scenarios, bridging the gap between empirical outcome modeling and mechanistic reasoning.

#### *S2. Data sources and standardization*

The PurpleLab claims database captures adjudicated medical and pharmacy claims across diverse payer types. The OMNY Health electronic health record (EHR) network aggregates structured clinical data from health systems across varied care settings and regions. Both databases provide geographic coverage across the United States and longitudinal follow-up between 2014 and 2025. Both data sources include diagnoses, procedures, medication exposures, and demographic information, with claims data providing comprehensive capture of healthcare utilization and dispensing events, and EHR data offering granular clinical detail, including laboratory results, physiological measurements, and documented clinical observations. Patients were de-identified and linked across sources by a third-party tokenization service (Datavant Inc., San Francisco, CA, USA).

All source data were standardized to the Observational Medical Outcomes Partnership (OMOP) Common Data Model to ensure consistent representation of clinical concepts, temporal relationships, and longitudinal patient histories across datasets. Standardized OMOP vocabularies were applied to harmonize conditions, drug exposures, procedures, and outcomes, enabling reproducible cohort definition and feature construction[1,2].

The drug-centric knowledge graph comprises approximately 150,000 biomedical entities and more than 7 million relationships spanning compounds, genes, proteins, diseases, pathways, and pharmacological attributes. It is constructed from over 30 curated biomedical databases, including DrugBank[3], ChEMBL[4], UniProt[5], and Reactome[6], and further enriched through semi-automated extraction from the biomedical literature with expert scientific curation. The graph encodes established relationships between drugs and entities such as diseases, molecular targets, mechanisms of action, signaling and metabolic pathways, and known drug–drug interactions. These relationships are used to derive drug-level representations (embeddings) that capture mechanistic and pharmacological similarity across therapies. Integration of these representations with longitudinal patient data enables the simulation framework to incorporate biologically informed structure while supporting generalization to therapies and contexts not directly observed in the underlying real-world data.

#### *S3. Construction of longitudinal patient histories*

For each patient, longitudinal health histories were constructed as time-ordered sequences of clinical events, beginning prior to cohort entry and extending through follow-up. Baseline covariates captured demographic characteristics, cardiovascular risk factors, comorbidities, prior cardiovascular events, and background therapies at cohort entry. Covariate trajectories were summarized using predefined windows, allowing the model to learn associations between evolving patient context and outcome risk. Feature definitions and temporal windows were fixed prior to prospective simulation to avoid outcome-driven refinement.

#### *S4. Outcome definition*

Cardiovascular outcomes were defined using prespecified code sets aligned with the primary endpoint definition for major adverse cardiovascular events used in the VESALIUS-CV trial, namely cardiovascular death, non-fatal myocardial infarction, and non-fatal stroke (3-point MACE), following the 2014 ACC/AHA key data elements and definitions for cardiovascular endpoint events[7]. Time to event was measured from the index date to the first occurrence of the outcome of interest or the end of available follow-up.

#### *S5. ClinBoost framework for cardiovascular time-to-event modeling*

Cardiovascular outcomes were modeled using the ClinBoost framework[8], a patient-level survival modeling approach designed to estimate time to first event from a defined index date. Each patient is assigned an index date representing trial entry, and the outcome label captures both event occurrence and time to event or censoring, enabling appropriate handling of right-censored data.

Patient representation incorporates structured clinical history prior to index and recent treatment exposure. Historical features are summarized over predefined time windows preceding index, while treatment is encoded based on unique drugs received within a fixed pre-index window.

ClinBoost uses an ensemble architecture combining gradient-boosted trees[9] and time-binned logistic regression layers[10] to estimate event probabilities across multiple time intervals. These interval-specific predictions are transformed into individualized survival curves, which form the basis for cumulative incidence estimation and downstream trial simulation.

#### *S6. Simulation cohort eligibility criteria*

The simulation cohort was defined to approximate a high cardiovascular risk primary prevention population with enriched metabolic and atherosclerotic burden. Inclusion criteria required evidence of at least one major cardiometabolic or vascular condition at baseline, including type 2 diabetes mellitus, peripheral vascular disease, established atherosclerotic cardiovascular disease, or ischemic cerebrovascular disease. In addition, patients were required to meet lipid-based risk thresholds, defined by elevated low-density lipoprotein (LDL) cholesterol (≥90 mg/dL) or non-high-density lipoprotein (non-HDL) cholesterol (≥120 mg/dL) within the 6 months prior to cohort entry.

Exclusion criteria were applied to remove patients with recent or unstable cardiovascular conditions and those with severe comorbidities that could confound outcome assessment. These included recent myocardial infarction, recent coronary artery bypass grafting, severe renal impairment (estimated glomerular filtration rate <15 mL/min/1.73 $m^2$), tachycardia, and severely reduced left ventricular ejection fraction (<30%). Patients with atrial fibrillation or flutter not receiving anticoagulation therapy were also excluded, as were those with markedly elevated triglycerides (≥500 mg/dL) and individuals undergoing recent or planned revascularization procedures.

Additional exclusions targeted recent acute cerebrovascular events and other high-risk clinical states to ensure a stable baseline population. Together, these criteria define a cohort enriched for cardiovascular risk factors and subclinical or stable disease, while minimizing confounding from acute events or advanced disease states, consistent with the intended population for cardiovascular outcomes trial simulation.

***S7. Virtual trial construction and generation of simulated trial outcomes***

To emulate the VESALIUS-CV trial (ClinicalTrials.gov NCT03872401), eligible patients were selected from real-world data using prespecified criteria aligned with the clinical protocol, and virtual trial cohorts were constructed to reflect the target population. For each patient, outcome trajectories were simulated under both investigational and comparator treatment scenarios, generating patient-specific counterfactual predictions for each arm. This approach enabled a balanced comparison of treatment effects independent of actual treatment exposure in the underlying real-world data. Allocation ratios and follow-up durations matched those of the target trial. All patients contributed risk information under their assigned virtual treatment arm for the duration of simulated follow-up or until event occurrence or censoring.

Patient-level predicted event-risk trajectories were aggregated within each arm to generate arm-level cumulative incidence functions. Cumulative probability curves were constructed from simulated time-to-event data to mirror the standard presentation of cardiovascular outcomes trials. These curves enabled visual and quantitative comparison of event accumulation and treatment effect emergence over time. Relative treatment effects were summarized using hazard ratios derived from simulated time-to-event data. All eligible patients contributed to estimation of the simulated event-risk trajectories under each virtual treatment scenario. Rather than selecting a subset of 6,150 patients per arm from the eligible patients pool, patient-level predictions from the full eligible cohort were aggregated to estimate the expected event rates over time for each treatment arm. These arm-level estimates were then used as the expected event probabilities for a simulated trial with a sample size of 6,150 patients per arm, matching the target trial design. Confidence intervals and P values were calculated based on this prespecified sample size and the corresponding simulated treatment-effect estimates. Follow-up duration and allocation ratio were aligned with those expected in the target trial. All trial-level estimates were generated and reported according to a standard analysis plan.

#### *S8. Post hoc population-matched sensitivity analysis*

To further assess the impact of population characteristics on simulated outcomes, a post hoc sensitivity analysis was conducted in which simulations were repeated using populations constructed to more closely match the baseline characteristics of the corresponding trial cohorts. For these analyses, the feature distributions of simulated patients were aligned to trial-reported characteristics, including demographics (age, sex, race, and ethnicity) and key clinical variables (hypertension, obesity, and diabetes), where available. Race and ethnicity were used as recorded in the source data, with the fixed categories of those sources. Results from these population-matched simulations were compared with primary simulation outputs to assess the sensitivity of both relative and absolute treatment effect estimates to population composition.

**Table S1.** Randomized cardiovascular outcomes trials used for trial-level model validation, with the reported statistical success of the investigational arm.

| Trial | ClinicalTrials.gov identifier | Investigational arm | Comparator | Phase |
|---|---|---|---|---|
| AMPLITUDE-O | NCT03496298 | Efpeglenatide | Placebo | 3 |
| CAMELLIA-TIMI 61 | NCT02019264 | Lorcaserin | Placebo | 3 |
| CANVAS | NCT01032629 | Canagliflozin | Placebo | 3 |
| CAROLINA | NCT01243424 | Linagliptin | Glimepiride | 3 |
| CLEAR Outcomes | NCT02993406 | Bempedoic acid | Placebo | 3 |
| CREDENCE | NCT02065791 | Canagliflozin | Placebo | 3 |
| DEVOTE | NCT01959529 | Insulin degludec | Insulin glargine | 3 |
| FLOW | NCT03819153 | Semaglutide | Placebo | 3 |
| FOURIER | NCT01764633 | Evolocumab | Placebo | 3 |
| Harmony Outcomes | NCT02465515 | Albiglutide | Placebo | 3 |
| LEADER | NCT01179048 | Liraglutide | Placebo | 3 |
| MK-3102-018 | NCT01703208 | Omarigliptin | Placebo | 3 |
| PIONEER 6 | NCT02692716 | Semaglutide (oral) | Placebo | 3 |
| REDUCE-IT | NCT01492361 | Icosapent ethyl | Placebo | 3 |
| REWIND | NCT01394952 | Dulaglutide | Placebo | 3 |
| SELECT | NCT03574597 | Semaglutide | Placebo | 3 |
| SOUL | NCT03914326 | Semaglutide (oral) | Placebo | 3 |
| SURPASS-CVOT | NCT04255433 | Tirzepatide | Dulaglutide | 3 |
| SUSTAIN 6 | NCT01720446 | Semaglutide | Placebo | 3 |
| TECOS | NCT00790205 | Sitagliptin | Placebo | 3 |
| THEMIS | NCT01991795 | Ticagrelor | Placebo | 3 |
| VERTIS CV | NCT01986881 | Ertugliflozin | Placebo | 3 |

[a]Success denotes a statistically significant benefit of the investigational arm on the primary cardiovascular endpoint (upper bound of the 95% confidence interval of the hazard ratio below 1).

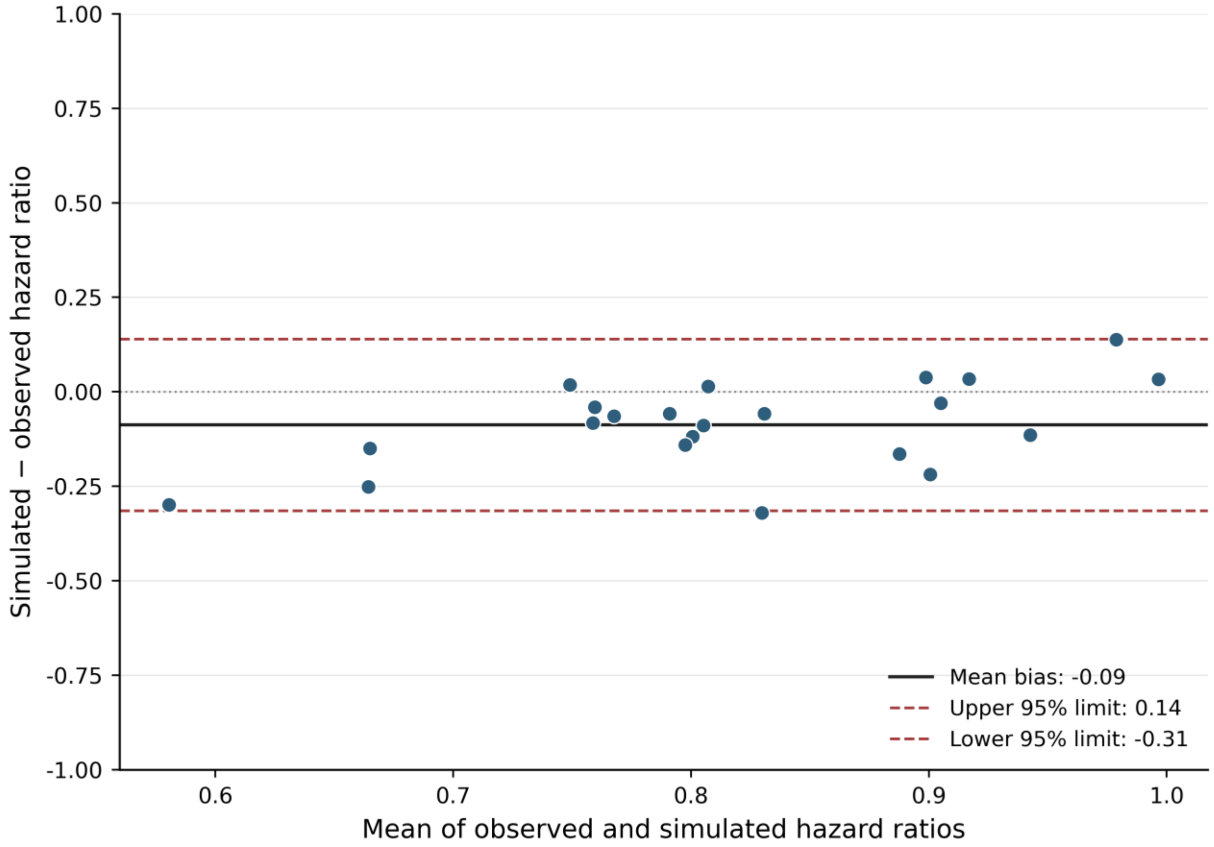


**Figure S1.** Bland–Altman analysis of observed versus simulated hazard ratios. Bland–Altman plot comparing hazard ratios between observed and simulated between-arm comparisons in the validation trials (n = 22). Red and blue dashed lines indicate the upper and lower limits of agreement (LOA); the grey dashed line represents the mean bias (-0.09; LOA −0.32 to 0.14).
